# Feasibility and acceptability for LION, a fully remote, randomized clinical trial within the VA for light therapy to improve sleep in Veterans with and without TBI: An MTBI^2^ sponsored protocol

**DOI:** 10.1101/2024.05.30.24308195

**Authors:** Jonathan E. Elliott, Jessica S. Brewer, Allison T. Keil, Brittany R. Ligman, Mohini D. Bryant-Ekstrand, Alisha A. McBride, Katherine Powers, Savanah J. Sicard, Elizabeth W. Twamley, Maya E. O’Neil, Andrea D. Hildebrand, Thuan Nguyen, Benjamin J. Morasco, Jessica M. Gill, Bradley A. Dengler, Miranda M. Lim

## Abstract

Sleep-wake disturbances frequently present in Veterans with mild traumatic brain injury (mTBI). These TBI-related sleep impairments confer significant burden and commonly exacerbate other functional impairments. Therapies to improve sleep following mTBI are limited and studies in Veterans are even more scarce. In our previous pilot work, morning bright light therapy (MBLT) was found to be a feasible behavioral sleep intervention in Veterans with a history of mTBI; however, this was single-arm, open-label, and non-randomized, and therefore was not intended to establish efficacy. The present study, LION (light vs ion therapy) extends this preliminary work as a fully powered, sham-controlled, participant-masked randomized controlled trial (NCT03968874), implemented as fully remote within the VA (target n=120 complete). Randomization at 2:1 allocation ratio to: 1) active: MBLT (n=80), and 2) sham: deactivated negative ion generator (n=40); each with identical engagement parameters (60-min duration; within 2-hrs of waking; daily over 28-day duration). Participant masking via deception balanced expectancy assumptions across arms. Outcome measures were assessed following a 14-day baseline (pre-intervention), following 28-days of device engagement (post-intervention), and 28-days after the post-intervention assessment (follow-up). Primary outcomes were sleep measures, including continuous wrist-based actigraphy, self-report, and daily sleep dairy entries. Secondary/exploratory outcomes included cognition, mood, quality of life, circadian rhythm via dim light melatonin onset, and biofluid-based biomarkers. Participant drop out occurred in <10% of those enrolled, incomplete/missing data was present in <15% of key outcome variables, and overall fidelity adherence to the intervention was >85%, collectively establishing feasibility and acceptability for MBLT in Veterans with mTBI.

## INTRODUCTION

Traumatic brain injury (TBI) can contribute to multiple short- and long-term negative neurologic sequelae, worsening functional outcomes and generating staggering economic burden [1]. One primary long-term complication associated with TBI (including mild TBI, mTBI) is sleep disturbance [2–4], with recent evidence demonstrating persistent post-concussive sleep-related impairment lasting several years [5] to >20 years post-injury [6–9]. The pathophysiology underlying persistent sleep disturbances after mTBI is unclear, but is potentially associated with damage to cortical pathways related to glutamate/GABA balance and/or orexin/hypocretin neurons, with subsequent effects on circadian-regulated systems such as melatonin and others [10–15]. Impaired sleep contributes to worse general health outcomes, including cognition, mood, and management of chronic pain, such that poor sleep exacerbates these outcomes which in turn further reduces sleep quality, creating a vicious cycle that is clinically very challenging to treat [16–21].

Clinically available interventions to treat sleep-wake disturbances, including in mTBI, often suffer from marginal efficacy, poor patient acceptability, and/or high patient/provider burden (e.g., medications, acupuncture, cognitive behavioral therapy for insomnia) [22–24]. Compounding this issue is the frequency of mTBI in Veterans and military service members, and associated high rates of mental health comorbidities, cognitive impairment, and chronic pain, all of which may limit the number of treatment options available or invite additional clinical considerations (e.g., pharmaceutical intervention, opioids, anti-depressants, or anxiolytics, to treat primary complaints, which often are accompanied by sleep impairing secondary effects including reducing slow-wave sleep, REM sleep or even melatonin production). Thus, alternative treatment approaches are needed in this population, particularly treatment options that are feasible, have high acceptability, are low burden, and have minimal side effects.

Light therapy, generally administered in the morning (i.e., morning bright light therapy; MBLT) is cost-effective, low-burden, home-based, and non-pharmacologic. Our recent single-arm, open-label clinical pilot study demonstrated preliminary feasibility and limited efficacy for MBLT to improve sleep, as measured both by self-report and objective actigraphy-based measures in Veterans with mTBI [25]. Much of the precedence for this pilot study stemmed from a large body of work associating mTBI with subsequent effects on sleep quality, mood and daytime alertness [26–39]. Additional rigorous studies have been executed comparing the effects of blue spectrum light with a placebo control amber light on sleep in individuals with mTBI, and found improvement in subjective daytime sleepiness, although objective measures of sleep via actigraphy were unchanged [40–42]. Two other recent studies examined light therapy on daytime fatigue in individuals with TBI (including severe TBI) [43,44]. Additionally, light therapy was also recently shown to improve post-traumatic stress disorder (PTSD) symptom severity in Veterans with PTSD, albeit without a change in self-reported or actigraphic metrics of sleep quality [45].

To our knowledge, the field of sleep medicine is without a fully powered, sham-controlled, masked, randomized controlled trial establishing feasibility, acceptability and efficacy of MBLT to improve sleep quality in Veterans with mTBI. Furthermore, to our knowledge, a fully remote randomized controlled trial device intervention protocol has not yet been successfully implemented in Veterans with mTBI – a challenging population not only due to significant participant disability/burden, but also due to the logistical and regulatory challenges of device management and electronic data capture behind the Veterans Affairs (VA) firewall. We sought to fill both gaps by implementing LION (Light vs Ion therapy), whose protocol is described herein. This trial proposed to acquire complete data in n=120 Veterans allocated 2:1, active-intervention (MBLT, n=80) versus sham-intervention (deactivated ion therapy, n=40). MBLT delivered white light (10,000 lux at the eye), whereas the negative ion generator was fully deactivated, but modified to appear “on”, i.e., via installation of a small fan to generate a quiet, whirring sound, as well as a small LED status light. This sham-intervention was modeled from prior work by Lam et al., [28] in which bright light or sham-intervention therapy was compared to fluoxetine for the treatment of depression. Participants in both groups engaged with devices to the same extent (i.e., over 60 minutes within 2 hours after waking) throughout the 28 day intervention period, with the same degree of expectancy and study team interaction. The brand of the light box was chosen due to being commercially available, in use for decades (including within the VA), and previously validated for illuminance metrics and spectral power density [25]. Dose, timing, and duration were all established from prior work investigating the effect of MBLT for the treatment of insomnia, circadian entrainment, and seasonal affective disorder [46–48].

The proposed sham-controlled, participant-masked, randomized controlled trial was fully powered to establish efficacy for MBLT to improve sleep in Veterans with mTBI. Furthermore, this study – innovative in that it is being implemented as a fully remote protocol within the VA – also provides valuable feasibility and acceptability outcomes for the remote aspects of the study design for future iterations. This trial is currently ongoing with ∼80% of participant enrollment completed. Therefore, this manuscript presents a detailed description of the clinical trial design, execution, interim feasibility/acceptability measures of the remote protocol, as well as the statistical analysis plan for future efficacy analyses once data collection is completed.

## MATERIALS & METHODS

### Overview & Ethics

LION is a sham-controlled, participant-masked, randomized controlled trial (NCT03968874), investigating primarily, the efficacy of morning bright light therapy (MBLT) to improve sleep quality, cognitive function, and quality of life in Veterans with mTBI, and secondarily, the feasibility and acceptability of implementing a fully remote clinical trial within the VA. Participants were randomized and allocated 2:1 to active (MBLT) or sham (negative ion generator) study arms, with a target total sample size of n=120 participants with complete pre- and post-intervention data (∼80% of the data has been collected so far). Primary outcomes were changes between groups from baseline to post-intervention (28 days), and baseline to follow-up (56 days). All participants that expressed interest, screened eligible, and were enrolled in the study were processed according to the Standard Protocol Items and Recommendations for Interventional Trials (SPIRIT) guidelines (**Figure 1**). This project is sponsored by the Military Traumatic Brain Injury Initiative (MTBI^2^) and approved by a joint Institutional Review Board (IRB) at the VA Portland Health Care System (VAPORHCS; #4268 and #4002) and Oregon Health & Science University (OHSU; #19411), as well as the Department of Defense Office of Human Research Oversight (OHRO). All participants provided, or will provide, verbal and written informed consent prior to participation.

**Figure 1.**
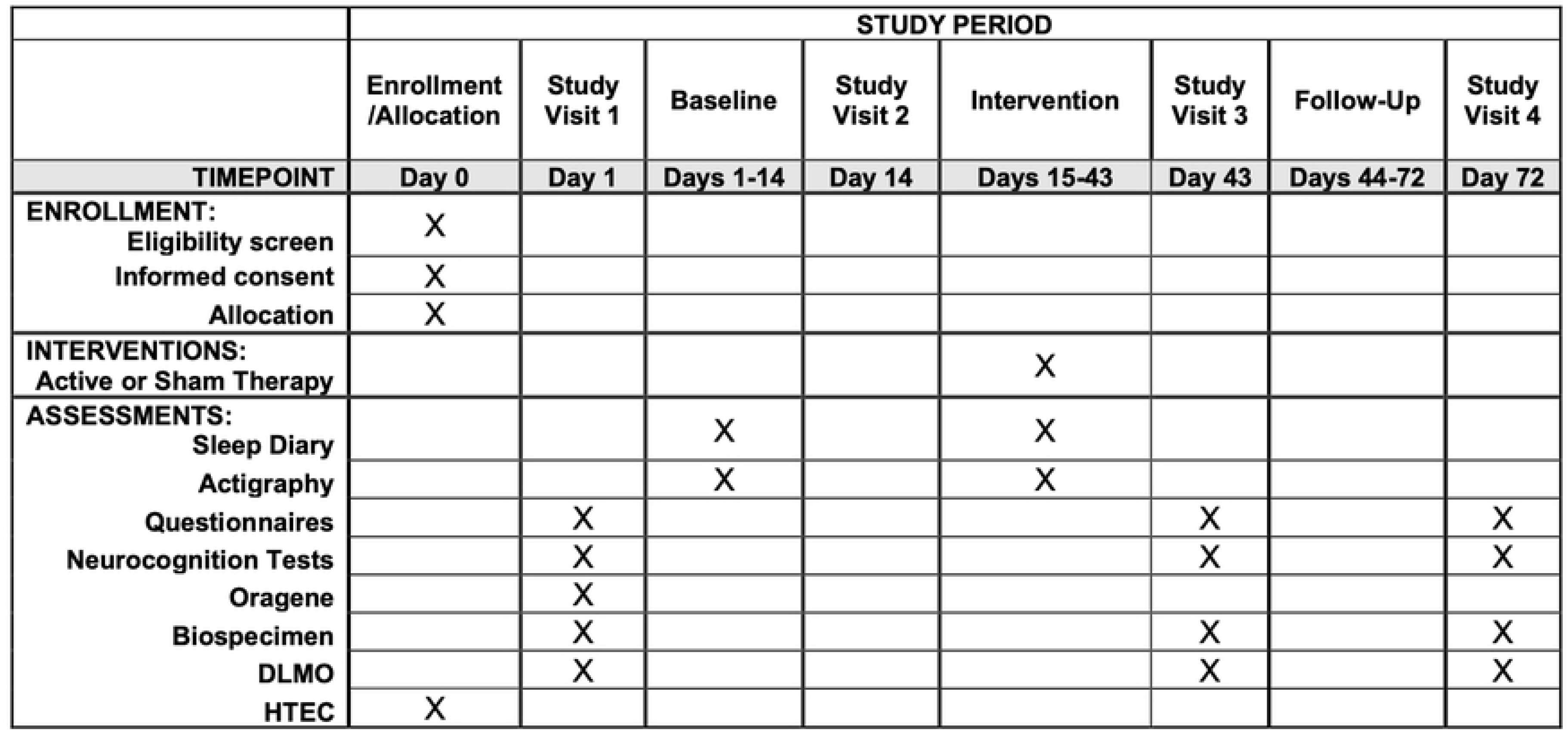
SPIRT outcomes across time. The Standard Protocol Items: Recommendations for Interventional Trials (SPIRIT) timeline for LION.

### Recruitment & Sample Selection

Participants were recruited nationwide, with additional local efforts from flyers and clinician referrals within the VAPORHCS, OHSU, and surrounding Portland-Metro area (**Figure 2**). The most successful recruitment methods included repositories, referrals, internet-based advertisements, flyers, and radio ads, all described in more detail below.

**Figure 2.**
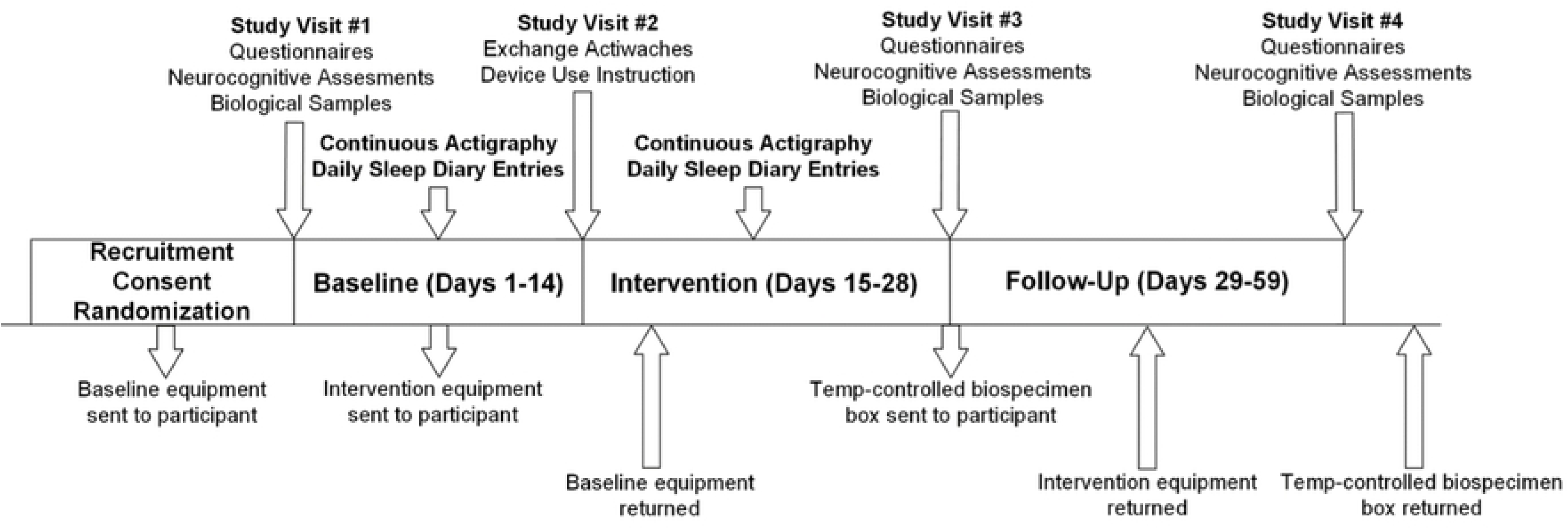
Participant flow chart. Diagram illustrating steps involved with LION, including when participant study visits are held, which outcomes are collected when, the duration of each study period as well as when equipment and biospecimens are shipped to/from participants place of residence.

#### Repositories

Several large participant repositories developed from prior and ongoing studies within the VAPORHCS were utilized, which collectively house contact information from over 1500 Veteran participants to date who consented to being recontacted for future research such as this study. Regulatory approvals were in place to share identifiable data and participant contact information with approved LION study team personnel. Additional tools that have or may be used in the future for participant recruitment include ResearchMatch.org and the VA Informatics and Computing Infrastructure (VINCI) system. Both services enable research coordinators to connect with potentially eligible participants either via personal profile information (research match) or through opt-out letters (VINCI).

#### Referrals

Several opportunities for participant referrals are approved and utilized. First, clinicians within the VAPORHCS and associated community-based outpatient clinics (including the university affiliate, OHSU) refer participants for further eligibility assessment. Second, ongoing studies within our research program at OHSU and the VAPORHCS (i.e., the Sleep & Health Applied Research Program [SHARP]) referred participants to the LION study who either did not qualify for other studies or completed those other studies. Third, MTBI^2^ provides a referral service for those interested in TBI research opportunities, called the TBI Research Opportunities and Outreach for Participation in Studies (TROOPS) program. In the case of TROOPS referrals, participant confidentiality in the communication between LION and TROOPS personnel is protected using a VA-authorized end-to-end encrypted email service for all messages containing PHI/PII.

#### Website Advertisement

The study utilized multiple venues for website advertisements. This included advertisements within the overall VAPORHCS website, as well as affiliated centers and labs (e.g., the VA National Center for Rehabilitative Auditory Research: www.ncrar.research.va.gov/Join_Research_Study/Index.asp). Similarly, the project is advertised on the OHSU research opportunity website, which is also shared with the OHSU Brain Institute (www.ohsu.edu/brain-institute/research-ohsu-brain-institute). Lastly, the study is listed on our research program’s website (www.sharplabpdx.com).

#### Flyers, Social Media, & Radio

Flyers were placed in high traffic areas of the main VAPORHCS hospital and at two outpatient clinics and shared with local clinicians, study sponsors and other partners for broad dissemination. A social media post for LION was placed on the MTBI^2^ blog that also contributed participants to the overall TROOPS referral program. Other approved social media venues included Craigslist advertisements, GovDelivery (a marketing subscription email service), and other VA-approved accounts. IRB-approved visual banners and audio ads that aired on local AM radio stations were also utilized to recruit participants within the Portland-metro area. See **Table 1** for the number of participants recruited via each approach.

**Table 1.** Interim demographic information for participants who have completed this protocol.

|  | **MBLT (n=69)** |  | **Sham (n=35)** |  |
| --- | --- | --- | --- | --- |
|  | mTBI (n=47) | No TBI (n=22) | mTBI (n=24) | No TBI (n=11) |
| **Age, years** | 53.2 ± 13.1 | 53.2 ± 14.8 | 50.9 ± 14.6 | 57.9 ± 15.9 |
| **Education, years** | 16.4 ± 1.9 | 15.8 ± 2.1 | 15.2 ± 1.9 | 16.2 ± 1.9 |
| **Veteran Status** |  |  |  |  |
| Veteran | 46 (98%) | 14 (64%) | 18 (75%) | 10 (91%) |
| Non-Veteran | 1 (2%) | 8 (36%) | 6 (25%) | 1 (9%) |
| **Gender** |  |  |  |  |
| Man | 31 (66%) | 11 (50%) | 17 (71%) | 4 (36%) |
| Woman | 15 (32%) | 11 (50%) | 7 (29%) | 7 (64%) |
| Non-binary/other | 1 (2%) | 0 (0%) | 0 (0%) | 0 (0%) |
| **Race and Ethnicity** |  |  |  |  |
| Ethnicity, *Hispanic/Latine* | 3 (6%) | 0 (0%) | 2 (8%) | 1 (9%) |
| Race, *White* | 37 (79%) | 14 (64%) | 17 (71%) | 11 (100%) |
| Race, *Asian* | 0 (0%) | 2 (9%) | 1 (4%) | 0 (0%) |
| Race, *Black or African American* | 1 (2%) | 4 (18%) | 0 (0%) | 0 (0%) |
| Race, *American Indian or Alaskan Native* | 5 (11%) | 0 (0%) | 4 (17%) | 0 (0%) |
| Race, *Two or More Races* | 2 (4%) | 1 (5%) | 1 (4%) | 0 (0%) |
| Race, *Other* | 1 (2%) | 1 (5%) | 1 (4%) | 0 (0%) |
| **Modality** |  |  |  |  |
| Remote (DocuSign consent) | 39 (83%) | 17 (77%) | 20 (83%) | 5 (45%) |
| In-Person (Physical copy consent) | 8 (17%) | 5 (23%) | 4 (17%) | 6 (55%) |
| **Participant Residence Time Zone** |  |  |  |  |
| Pacific Standard Time | 30 (64%) | 17 (77%) | 20 (83%) | 10 (91%) |
| Local Portland-Vancouver metro area | 15 (32%) | 8 (36%) | 9 (38%) | 5 (45%) |
| Mountain Standard Time | 3 (6%) | 0 (0%) | 0 (0%) | 0 (0%) |
| Central Standard Time | 4 (9%) | 1 (5%) | 2 (8%) | 0 (0%) |
| Eastern Standard Time | 8 (17%) | 4 (18%) | 1 (4%) | 1 (9%) |
| **Recruitment Method** |  |  |  |  |
| Repository | 3 (6%) | 1 (5%) | 5 (21%) | 3 (27%) |
| Referral | 11 (23%) | 5 (23%) | 1 (4%) | 2 (18%) |
| Website | 5 (11%) | 5 (23%) | 2 (8%) | 0 (0%) |
| Flyers, social media, and radio | 27 (57%) | 10 (45%) | 16 (67%) | 4 (36%) |

Data are presented as mean ± standard deviation, or *n* (%).

### Eligibility Criteria

Eligibility criteria for enrollment included, over 18 years of age, able to provide informed consent and comply with the study protocol, no history of macular degeneration (contraindicated due to light therapy potentially exacerbating underlying pathology), no history of bipolar disorder (contraindicated due to light therapy potentially inciting a manic/depressive episode [31]), no shift work on third shift (e.g., 2300 – 0700 or equivalent) or any amount of days overnight, no current engagement in light therapy, and stable status on other pharmacological and/or behavioral sleep interventions.

This study originated in an in-person format with an exclusive focus on Veterans with mTBI, however protocol #4268 was amended to be conducted remotely, and expand recruitment to non-Veterans and to non-TBI participants, for the express purpose of increasing the diversity of this sample in terms of the number of women and individuals from racial/ethnic minority groups and increasing overall generalizability. See **Table 1** for demographic parameters describing this current interim analysis.

History of mTBI was confirmed through a structured clinical interview using the Head Trauma Events Characteristics (HTEC; **Table 2**). The HTEC consists of a standard screening question followed by branching logic questions addressing injury type, location, intracranial injury/skull fracture, duration of loss of consciousness (LOC), and anterograde or retrograde post-traumatic amnesia (PTA), conferring a diagnosis of no, mild, moderate or severe TBI. Participants with an affirmative TBI outcome on the HTEC were excluded if their injury was moderate to severe and thus, the study only included participants with a history of mTBI. However, participants screening negative for TBI on the HTEC who met all other eligibility criteria, were also offered enrollment to promote generalizability throughout the wider VA.

**Table 2.**
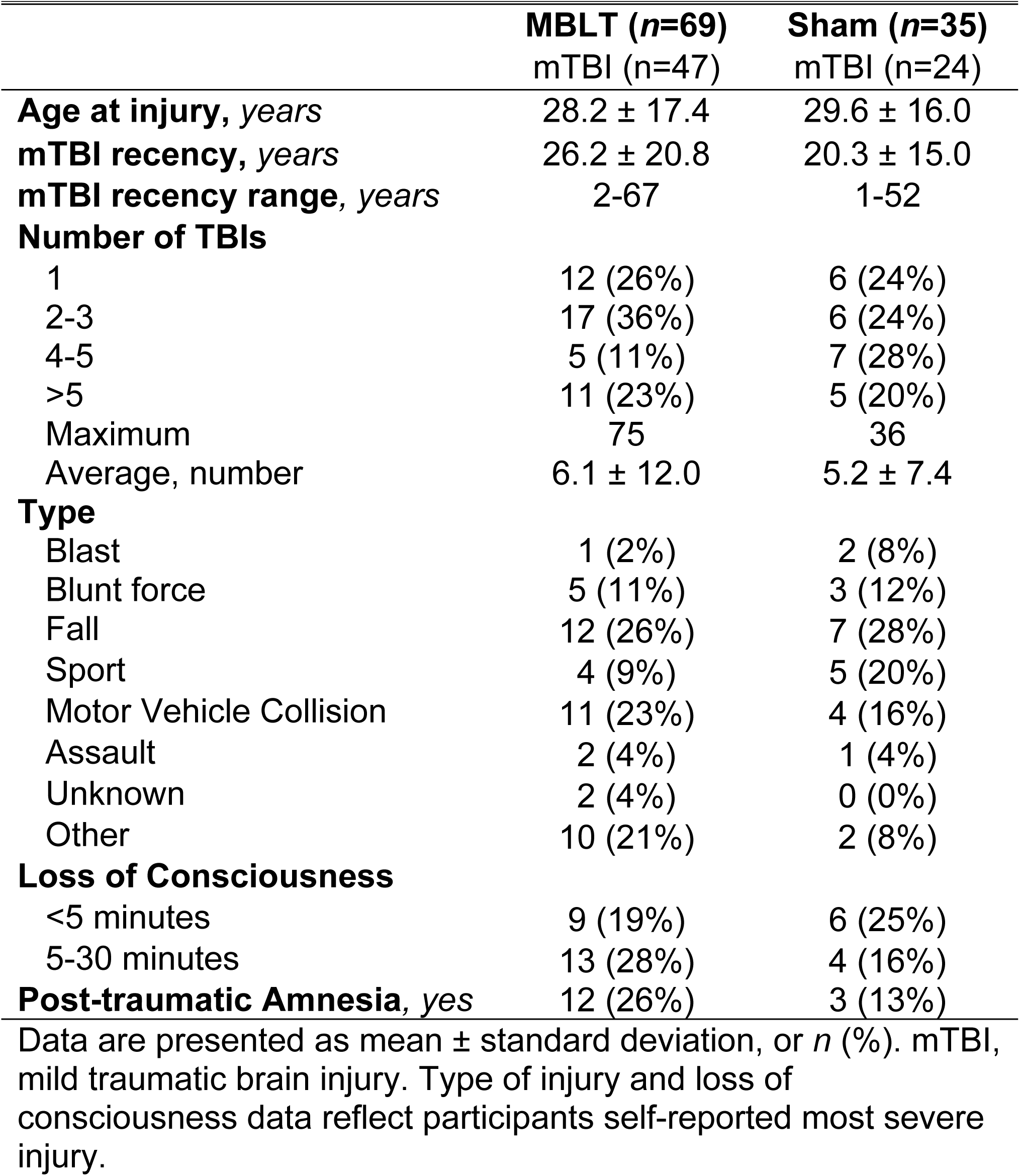
Current HTEC derived mTBI metrics.

**Table 3.** Feasibility and acceptability.

|  | **MBLT (n=69)** |  | **Sham (n=35)** |  |
| --- | --- | --- | --- | --- |
|  | mTBI (n=47) | No TBI (n=22) | mTBI (n=24) | No TBI (n=11) |
| **Device Adherence** |  |  |  |  |
| Self-reported use, *days* | 22.8 ± 7.7 (82%) | 24.6 ± 6.4 (88%) | 22.1 ± 8.5 (79%) | 23.5 ± 6.5 (84%) |
| Objective estimate, *days* | 22.4 ± 7.7 (80%) | 22.4 ± 8.2 (80%) | - | - |
| **Primary Sleep Outcome** |  |  |  |  |
| <u>Insomnia severity index</u> |  |  |  |  |
| Pre-intervention | 47 (100%) | 22 (100%) | 24 (100%) | 11 (100%) |
| Post-intervention | 43 (91%) | 17 (77%) | 21 (88%) | 10 (91%) |
| **Additional Sleep Outcomes** |  |  |  |  |
| <u>Actigraphy</u> |  |  |  |  |
| Pre-intervention, *days* | 12.4 ± 3.5 (89%) | 12.3 ± 0.2 (88%) | 11.2 ± 3.3 (80%) | 13.5 ± 5.4 (97%) |
| Post-intervention, *days* | 23.5 ± 7.8 (84%) | 25.6 ± 6.2 (91%) | 24.9 ± 8.2 (89%) | 23.8 ± 6.9 (85%) |
| <u>Sleep Diary</u> |  |  |  |  |
| Pre-intervention, *days* | 13.0 ± 3.4 (93%) | 14.0 ± 3.4 (100%) | 13.0 ± 4.9 (93%) | 11.0 ± 0.5 (79%) |
| Post-intervention, *days* | 24.9 ± 8.3 (89%) | 25.5 ± 6.4 (91%) | 24.8 ± 8.2 (89%) | 25.0 ± 6.2 (89%) |
| **Secondary Outcomes** |  |  |  |  |
| <u>Full questionnaire battery</u> |  |  |  |  |
| Pre-intervention | 47 (100%) | 22 (100%) | 24 (100%) | 11 (100%) |
| Post-intervention | 43 (91%) | 17 (77%) | 20 (83%) | 11 (100%) |
| <u>Cognitive Assessment</u> |  |  |  |  |
| Pre-intervention | 47 (100%) | 22 (100%) | 24 (100%) | 11 (100%) |
| Post-intervention | 45 (96%) | 22 (100%) | 23 (96%) | 11 (100%) |
| <u>DLMO</u> |  |  |  |  |
| Pre-intervention | 40 (85%) | 18 (82%) | 21 (88%) | 10 (91%) |
| Post-intervention | 38 (81%) | 19 (86%) | 20 (83%) | 11 (100%) |
| <u>Biospecimen sample</u> |  |  |  |  |
| Pre-intervention | 41 (87%) | 19 (86%) | 21 (88%) | 9 (82%) |
| Post-intervention | 40 (85%) | 17 (77%) | 20 (83%) | 6 (55%) |

Data are presented as mean ± standard deviation, or *n* (%). Pre-intervention assessments span a maximum of 14 days. Post-intervention assessments and device adherence spans a maximum of 28 days. DLMO, dim light melatonin onset.

### Screening & Informed Consent

Eligibility screening occurred either in-person or over the phone following an IRB-approved script. Depending on the route of recruitment, some participants completed a “pre-screening” online survey via the Research Electronic Data Capture (REDCap) system. Any participants who completed this pre-screening survey received a separate phone call to complete screening prior to advancing to informed consent.

Informed consent was completed over the phone or via video conference with the participant and a study coordinator according to standard practice, which included an addendum for permission to store their data and biological samples for future analyses and studies. Consent was obtained either via physical mail of consent forms to participants’ residences together with a live phone call, or, alternatively, electronically via VA-approved DocuSign (www.docusign.com). DocuSign is an e-signature service that hosts this study’s informed consent document and HIPAA authorization for digital signatures – enabling same-day screening and consenting – and was new to the VA system as of March 2021. Digital delivery of documentation was sent using DocuSign’s encryption service with a fully signed copy sent to both the study participant and research team. Local storage of this documentation was housed on the secure, firewall-protected, VA Research Drive. Significant advantages existed using DocuSign versus physically mailing documentation back and forth, including a substantial time savings and minimization of errors (e.g., missing a signature). One caveat was that DocuSign did not adjust times when signing parties were in different time zones; therefore, in these cases, an additional Note to File was included to explain any large discrepancies in timestamps between consenters and participants.

### Randomization, Allocation, & Masking

After receiving informed consent, HIPAA authorization, and completing the 2-week baseline period, participants were randomized in a 2:1 allocation ratio, active intervention to sham and recorded according to the Consolidated Standards of Reporting Trials (CONSORT; **Figure 3**). Although participant-masked (i.e., investigators know all negative ion generators are inactive), study arms were verbally described to participants in a “pseudo” double-masked fashion, as before [28]. Specifically, all participants were informed that 50% of both study arm devices were inactive and neither they, nor us, knew whether they receive an active device until completion. After the study had been completed, a team member then disclosed to all participants their randomization status, and provided the scientific rationale necessitating the use of deception. Additionally, study investigators described these study arms as “devices”, rather than “therapies”, throughout the protocol, which was technically accurate and precluded unmasking.

**Figure 3.**
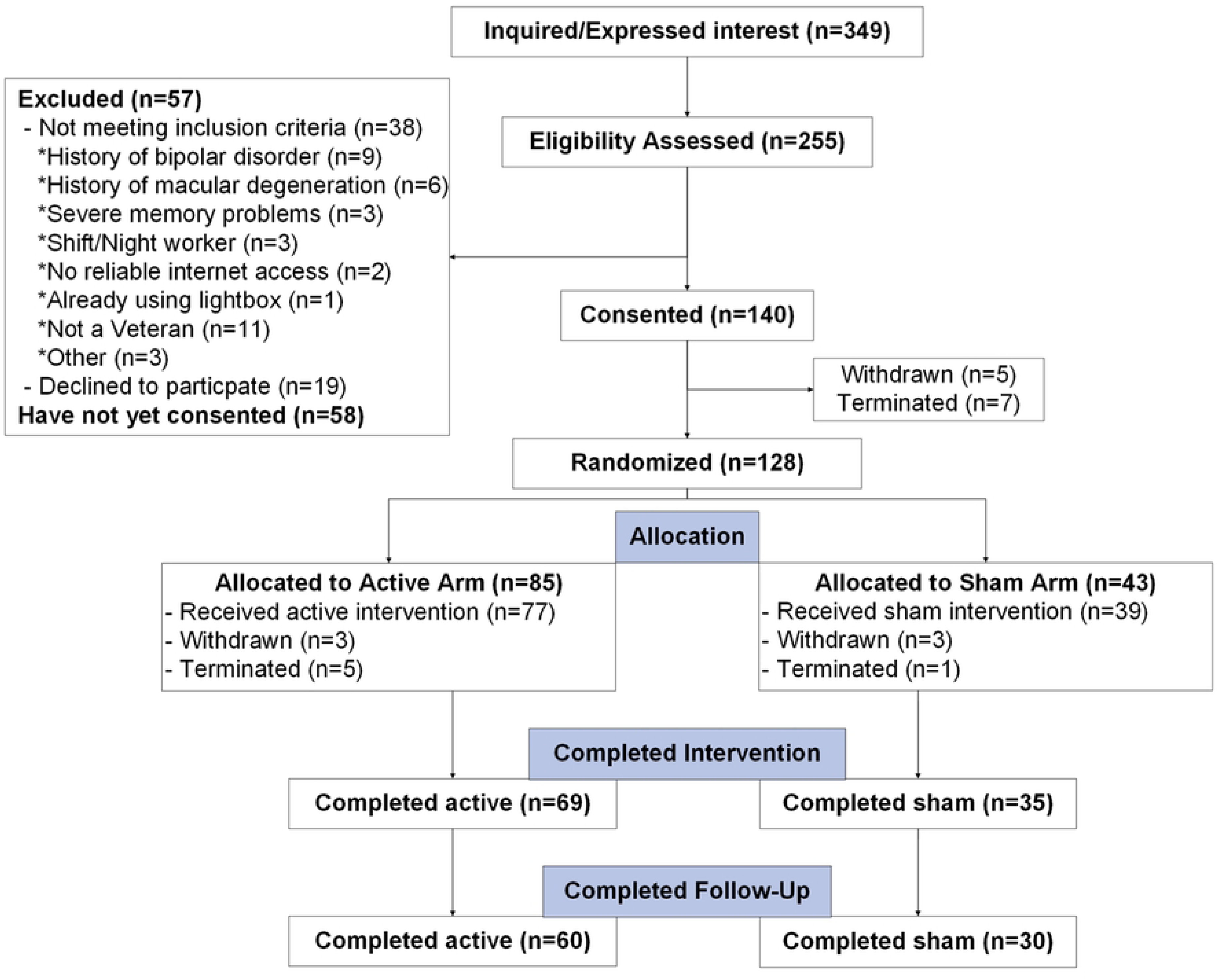
CONSORT Diagram. The Consolidated Standards of Reporting Trails (CONSORT) diagram for participants in LION to date.

### Interventions

#### Active – Morning Bright Light Therapy (MBLT)

Participants randomized to receive MBLT were shipped a light box (LightPad, Aurora Light Solutions, Las Vegas, NV, USA) to be received following the end of their baseline period, as we have previously published [25]. Participants were instructed on how to operate the LightPad, which involved plugging it in and pressing the “on” button. However, the lux received depends on the distance participants sit from the light source and thus, participants were instructed to sit no less than 24 inches away to ensure exposure to 10,000 lux of non-UV light. This was the maximum illuminance/range in commercially available units, and the recommended illuminance for white light to promote synchronization of circadian sleep-wake cycles in humans [29]. Participants were instructed to obtain their light exposure within 120 minutes of waking. Thus, participants were allowed 60 minutes of flexibility in their morning routine before MBLT, while still receiving 60 minutes of bright light within the advised post-wake window. Detailed instructions, including visual aids, were provided to promote compliance (e.g., positioning the light box at a 45° angle relative to their face), noting that instructions for light therapy had previously been shown to be significant moderators of efficacy [49]. Participants were also encouraged to concurrently engage in other normal activities of daily living during this time (e.g., read a newspaper, work on a computer, eat breakfast).

#### Sham – Deactivated Negative Ion Generator

The sham intervention consisted of a no-light, negative ion generator (SphereOne, Inc.) of comparable size and electricity demands to the LightPad, also shipped to participants at the end of their baseline period. All negative ion generator devices were modified to emit an audible quiet hum with a green status light, and deactivated to not emit ions, as previously published [28], which falsely indicated being in an active state. The sham arm followed identical timing and duration as described above for the active-MBLT arm. Therefore, participants in the sham arm also received the same degree of subject-investigator interaction and instructions for device adherence.

#### Fidelity Adherence Monitoring

Adherence to both MBLT and sham was tracked and consistently monitored in several ways. First, we called participants at the onset of initiating their intervention, as well as weekly thereafter to address any questions and promote their adherence by way of addressing any challenges they had encountered. Second, participants manually recorded their engagement with their device (time and duration) as part of their daily sleep-diary. Third, we utilized a combination of actigraphy and luxometer data (generated and collected by the Actiwatch-2 as well as a HOBO Pendant MX Temperature/Light Data Logger affixed to the LightPad) to objectively track MBLT adherence. Luxometer data via the HOBO and/or actiwatch provided evidence of device usage, and activity counts via the actiwatch provided an estimate of movement such that the combination of low activity counts (i.e., suggesting being sedentary) and evidence for device usage, suggested protocol adherence. This did not preclude the possibility that participants turn the LightPad on and either have it incorrectly placed (facing away from them), or simply remain sedentary in a different room, but the combination of these metrics provided a best approximation while minimizing participant burden.

### Outcomes

#### Primary

LION’s primary outcome was designated as an improvement in sleep following MBLT compared to sham. This was accomplished via several means, though self-reported sleep disturbances via the Insomnia Severity Index (ISI) remained primary [50]. The ISI is a 7-item measure, each item a 5-point Likert scale (0-4) with a total score range of 0-28 where higher scores indicate worse sleep [51,52].

#### Self-report, actigraphy, and sleep diary

Potential changes in sleep were further evaluated via other validated self-report measures (e.g., Epworth Sleepiness Scale, ESS [53]; Functional Outcomes of Sleep Questionnaire-10, FOSQ-10 [54]; and the Sleep Hygiene Index, SHI [55]), and objective metrics derived from participants’ sleep diary and wrist-based actigraphy (Philips Respironics, Bend, OR, USA). Philips Actigraphy devices use a solid-state, piezoelectric, monoaxial accelerometer to measure activity and silicon photodiodes to measure photopic light (5-100,000 lux; 400-900 nm), with data sampled at 32 Hz and aggregated into 2-minute bins.

Philips Actiware (v 6.3.0) was used to process and analyze these data relying first on the automated algorithmic detection of rest-active periods, with further refinement based on *a priori* defined outcomes collected in the sleep diary. For example, participants were queried whether their sleep was unusually disrupted (ranging from being ill to falling asleep or being awoken at unusual times due to things outside of their control), and if so, this 24 hour period was excluded from analysis. Participants were directed to wear the watch on their non-dominant wrist and to keep clothing from covering the light sensor as much as possible.

Daily sleep diaries were collected using the TWILIO™ platform, which directly integrated with our HIPAA-compliant secure REDCap database. Participants were prompted in the morning via Short Message Service (SMS) text format. The first prompt was at 8:00 AM in participants local time, with reminders sent at 10:00 AM and 12:00 PM if no response had been entered. If participants did not respond within that 24 hour period, earlier prompts expired ensuring all data is collected within 24 hours. Sleep diary metrics included self-reported time in bed/asleep, time out of bed/awake, number of awakenings during the night, and (during the intervention period) device usage.

The primary comparison across timepoints was pre-vs post-intervention. Only self-reported sleep symptoms were evaluated at the follow-up time point as daily diaries and actigraphy were not continued during the follow-up period.

#### Secondary Self-report

Additional relevant outcomes included a broad range of validated self-reported outcomes related to neurobehavioral function, mood, and quality of life (e.g., NIH PROMIS Global Health, Pain Intensity/Interference, and Emotional Distress Anxiety [56] [57]; Neurobehavioral Symptom Inventory, NSI [58]; Post-traumatic Stress Disorder Checklist for DSM-5, PCL-5 [59]; Patient Health Questionniare-9, PHQ-9 [60]; World Health Organization Disability Schedule 2.0, WHO-DAS 2.0 [61]; and Sleep Hygiene Index, SHI [55]). On average, this questionnaire battery required 30-45 minutes to complete. Questionnaires were completed using REDCap, with all entries automatically recorded. If participants were unable to complete questionnaires electronically, these were printed, mailed and all entries manually recorded into REDCap with a secondary rater confirming data entry accuracy.

#### Secondary: Cognition

Neurocognitive outcomes were assessed with both subjective and objective measures, and evaluated through neuropsychological testing (pre-, post-, and follow-up). This assessment was ∼30 minutes in length and completed during a video call evaluating memory, attention, executive function, language and processing speed (e.g., Delis-Kaplan Executive Function System (D-KEFS) Verbal Fluency [62]; Controlled Oral Word Association Test, COWAT-FAS [63]; Wechsler Adult Intelligence Scale, Fourth Edition, WAIS-IV, Digit Span Arithmetic subtest [64]; Hopkins Verbal Learning Test – Revised, HVLT-R ). The HVLT-R implemented Form 1 pre-intervention and on follow-up, with Form 4 for post-intervention. This evaluation was also recorded for the accurate transcription of participant responses. [65]

#### Secondary: Biomarkers

This protocol evaluated biofluid-based biomarkers associated with changes in sleep quality. Prior to the COVID-19 pandemic, this was accomplished through blood draws with appropriate processing to store serum/plasma aliquots (the first n=30 participants). However, during the months of March 2020 and onward, the study pivoted to a fully remote protocol and as such, implemented sweat patches (PharmChek, Fort Worth, TX, USA) as a means to assay a comparable scope of blood-based biomarkers that were implicated in our prior pilot study on light therapy and sleep in TBI [25]. Assays included markers of neuroinflammation and degeneration (e.g., Neurofilament Light Chain, NfL; Glial Fibrillary Acidic Protein, GFAP; Ubiquitin Carboxyl-Terminal Hydrolase L1, UCH-L1; and total Tau, t-Tau), as well as markers of systemic inflammation (e.g., Interleukin-6, IL-6; Interleukin-10, IL-10; and Tumor Necrosis Factor-alpha, TNF-α). A small internal validation study was conducted comparing body locations as well as duration of affixing sweat patches that informed the current protocol as follows (data not shown): Participants were instructed to affix the sweat patch on their lower abdomen, after cleaning the area with an alcohol swab, and to keep the sweat patch on for 24 hours. Once the 24-hour period was complete, the sweat patch was removed, placed in a labeled plastic bag, and stored in the participant’s freezer. All sweat patches (pre-, post-, and follow-up) remained in the participant’s freezer until the end of the follow-up period, when they were provided with a pre-paid FedEx cold-shipping box (FedEx Nanocool) maintaining an internal temperature of ∼4C for up to 96 hours to send the samples back to the study site. Upon receipt, the internal temperature was confirmed and sweat patches were stored at -20C until ready for batch assays.

#### Secondary: Dim Light Melatonin Onset

The protocol instructed participants to complete a salivary Dim Light Melatonin Onset (DLMO) protocol for the evaluation of changes in circadian alignment [66,67]. Detailed instructional guides reviewing the protocol were provided, a pair of blue wavelength blocking glasses, a toothbrush, and seven cotton-filled salivettes were provided. Samples were collected hourly for 5 hours prior to habitual bedtime, at bedtime, and one-hour post-habitual bedtime (i.e., participants stayed awake for 1 hour later than their habitual bedtime to provide 7 samples across 7 hours). Participants were advised to shutter blinds and turn off/dim lights while also wearing blue wavelength blocking glasses throughout the duration of this assessment, with objective light exposure also captured via the actiwatch’s luxometer. Also, participants were asked to refrain from alcohol for 72 hours before collection, and before each sample, remain seated and lightly brush/rinse their mouth with water (toothpaste/mouthwash is not allowed). Each sample was labeled and stored in the participants freezer until conclusion of follow-up whereby all three sets of samples were returned via the aforementioned temperature-controlled shipping container, following triple containment guidelines set by the Department of Transportation for biological samples. Upon receipt, internal temperature was confirmed and DLMO samples were processed according to standard procedures, aliquoted, and stored at -80C.

#### Secondary Genetic analyses

Lastly, the protocol included the collection of an Oragene DNA sample (DNAgenotek, Ottawa, Ontario, CA) that consisted of ∼2 ml of saliva. Participants avoided eating and drinking for 30 minutes before collection. Samples were considered stable at room temperature and therefore returned when convenient, generally occurring when participants returned their baseline period actiwatch.

### Shipping

The United States Postal Service (USPS) served as the primary courier for LION study packages to and from the participant’s home (**Figure 2**). The protocol specified that USPS flat rate boxes would be sent before baseline and intervention periods that contain all necessary instructions and equipment. Pre-paid return labels were already affixed underneath the primary delivery label. Unforeseen weather or other events preventing USPS from delivering these shipments on time was monitored and research coordinators rescheduled study visits if needed. Delivery of each shipment occurred before the baseline visit and before the onset of intervention (which includes randomized study arm device), with equipment returned using the same physical box it was received in. If needed, replacement return boxes were sent. Finally, at the end of the follow-up period, study coordinators sent the aforementioned FedEx cold-shipping box to return biospecimens. Receipt of returned equipment occurred following baseline and intervention. Study coordinators sanitized actiwatches with a 10% bleach solution followed by a 70% isopropyl alcohol solution, and downloaded the data to our Philips Actiware database after which it was charged and reconfigured for a new participant. Intervention devices were similarly sanitized and repackaged into protective storage containers.

### Data Safety, Monitoring & Auditing

#### Study Sponsor Monitoring

The study sponsor, MTBI^2^, was responsible for maintaining quality assurance and accuracy of data collection, approved modifications, reviewed secondarily to our local IRB, and monitored our study site and study personnel to ensure ethical research conduct. The sponsor conducted in-person or remote monitoring visits twice per year, in which a study monitor from MTBI^2^ reviewed deidentified data, required reporting documents and the timeline of the study. Deidentified data and regulatory documents were stored on MTBI^2^’s data collection website, Collection Access Sharing Analytics (CASA), and hosted at the NIH for ease of access to monitors. A physical copy of regulatory documents was maintained at the site location. Study coordinators used the resources on MTBI^2^’s CASA to generate Globally Unique Identifiers (GUIDs) for the participants (using participant’s full name, date of birth, city they were born in); a master key list was maintained by study personnel and coded data are kept separate from personal health information.

#### OHSU Oregon Clinical and Translational Research Institute

OCTRI hosted and managed data security and backup of the HIPAA-compliant REDCap database where our participant response data were stored. They have checks and balances coded into the database to allow for proper data protection, historical information and qualitative notes to be maintained. Access was protected via OHSU login information and user credentials once the database was shared from previously approved study personnel. This database is HIPAA-secure and all those interacting with the database are trained and knowledgeable of HIPAA and VA privacy policy.

### Statistical Analyses

The statistical analysis plan was designed to be carried out using GraphPad Prism v9 or R, with alpha defined *a priori* at 0.05, using a thorough descriptive analysis of participant baseline characteristics prior to evaluation of outcomes. Categorical variables will be described using frequencies and percentages. Histograms and boxplots will be used to assess the distribution of continuous variables. Continuous variables that follow an approximately normal distribution will be summarized using means and standard deviations; skewed continuous variables will be reported as medians and interquartile ranges. Potential covariates will also be summarized with descriptive statistics and graphs. As part of our descriptive analyses, we will compare participants who complete versus drop out of the study by baseline characteristics and study arm to ascertain potential biases that may impact this and future studies. Effect size estimates will be reported as Cohen’s d (small = 0.2; medium = 0.5; large = 0.8) or standardized regression coefficients (β) (small = 0.14; medium = 0.39; large = 0.59). We will also report the number needed to treat (NNT) approximated using methods described by Kraemer and Kupfer [68]. The Benjamini-Hochberg procedure will be used to control the false discovery rate for primary outcomes. Statistical outliers will be identified using regression diagnostics to estimate Cook’s Distance, and then a cutoff of 4/n (n = number of participants) will be used to identify outliers. Outliers will be examined for data entry errors, but otherwise retained in analyses. If data violate parametric assumptions via Shapiro-Wilk test, test statistics will be derived using robust standard errors. We will use the intent-to-treat principle and include all participants randomized to treatment irrespective of treatment compliance.

To analyze treatment effect over time (pre-, post-, and follow-up), we will implement mixed effects models (either linear or gamma, pending data normality) for each outcome measure under consideration. This flexible approach allows us to examine the change pattern and accounts for within-subject correlations over multiple time points. This approach also maximizes our observed data by way of accommodating missing data at random. The main effects in each model will be the treatment and time effects, including a treatment-by-time interaction effect. The models may be adjusted for possible confounders, such as age, sex, fidelity adherence (lux/min), geographic location/seasonality, sleep hygiene, and depression. With a significant omnibus test suggesting the global null hypothesis is rejected, a Bonferroni or Tukey HSD post-hoc analysis will be performed.

Within the MBLT group we will use linear regression to analyze the relationship (goodness of fit; r^2^) between average morning lux exposure (quantified as lux/min by the Actiwatch) and the degree of change in dependent variables (e.g., sleep efficiency, self-reported sleep quality, neurocognitive outcomes, and other self-reported functional outcomes) between pre- and post-treatment. Within select patient outcome questionnaires, we will perform Pearson correlational analyses to determine how each patient outcome questionnaire varies together (Pearson correlation coefficient; r). Actigraphy variables will be compared to appropriate self-report and objective outcomes in the form of discrete time-period averages (i.e., 3-5 consecutive representative nights within the last ∼7 days of both the baseline and intervention periods).

Secondarily, if permissible, we will implement analyses based on a 3-day moving average, leveraging the unique frequency of data acquisition inherent to 24/7 actigraphy.

We will examine patterns of missingness in outcomes over time. Every effort will be made to minimize the risk of systematic missing data. Sensitivity analyses will be performed to assess the influence of missingness and attribute this to either “Random” or “System” causes (appreciating that data are rarely missing completely at random). These analyses will inform the robustness of our findings and potentially obtain better estimates of the magnitude of effects to inform future trials. We will follow the recommendations by Jakobsen et al.[69] to handle missing data for our assessment: (1) <5% without evidence of differential patterns of missingness we will consider this negligible and impute the “best” and “worst” case scenarios for missing data to estimate plausible ranges of MBLT and placebo effects. (2) 5-25% we cue implementation of Markov Chain Monte Carlo (MCMC) multiple imputation to impute missing values and analyze 5 combined imputed datasets. (3) >25% (unlikely) we will not attempt to impute missing values. In the case of (3), the results of the complete case analyses will be reported along with reasons for missingness. In all cases, we will discuss and report the extent of missingness with a clear discussion of study limitations.

## RESULTS

Formal statistical analyses across participant groups (**Tables 1 and 2**) or in primary/secondary outcomes have not yet been explored given the ongoing/incomplete status of this randomized controlled trial. However, a central goal of presenting this study protocol was to evaluate overall feasibility and acceptability. This was best demonstrated through evaluating overall participant retention (feasibility; **Figure 3**) and a combination of protocol fidelity adherence and “data completeness” for primary

With respect to participant retention, of the *n*=140 participants who have provided informed consent, *n*=24 have withdrawn or been terminated (**Figure 3**), and therefore *n*=116 (83%) have been retained. At the time of this interim analysis, complete pre- and post-intervention data has been recorded in n=104 participants and therefore the remaining n=12 participants have been randomized/allocated and are currently ongoing.

In establishing protocol acceptability, we first considered the rate of missing data and other errors present in major outcome assessments, within the 14-day baseline period. This was examined via participant response rates for sleep diary entries and actigraphy usage (**Figure 4A and 4B**). Response rates for daily sleep diary entries did not differ between the active (97%) and sham (86%) conditions (*p*>0.05). Similarly, the number of days participants provided usable actigraphy data did not differ between the active (91%) and sham (89%) conditions (*p*>0.05). Protocol acceptability specific to the intervention examined overall fidelity adherence, which has averaged 83-93% across study arms. No differences were detected in the rates of participants meeting threshold for fidelity adherence across study arms when referencing either self-report or the objective assessment (*p*>0.05). Noting that there was not an objective assessment of device adherence to the sham condition. This protocol’s *a priori* definition of “fully adherent” was that participants engage with the device on at least 70% of the intervention period (5 out of every 7 days; therefore, a minimum total of 20 out of 28 days). Accordingly, device adherence has on average exceeded expectations with most adherent participants reporting device use for >23/28 days (**Figure 5A and 5B**).

**Figure 4.**
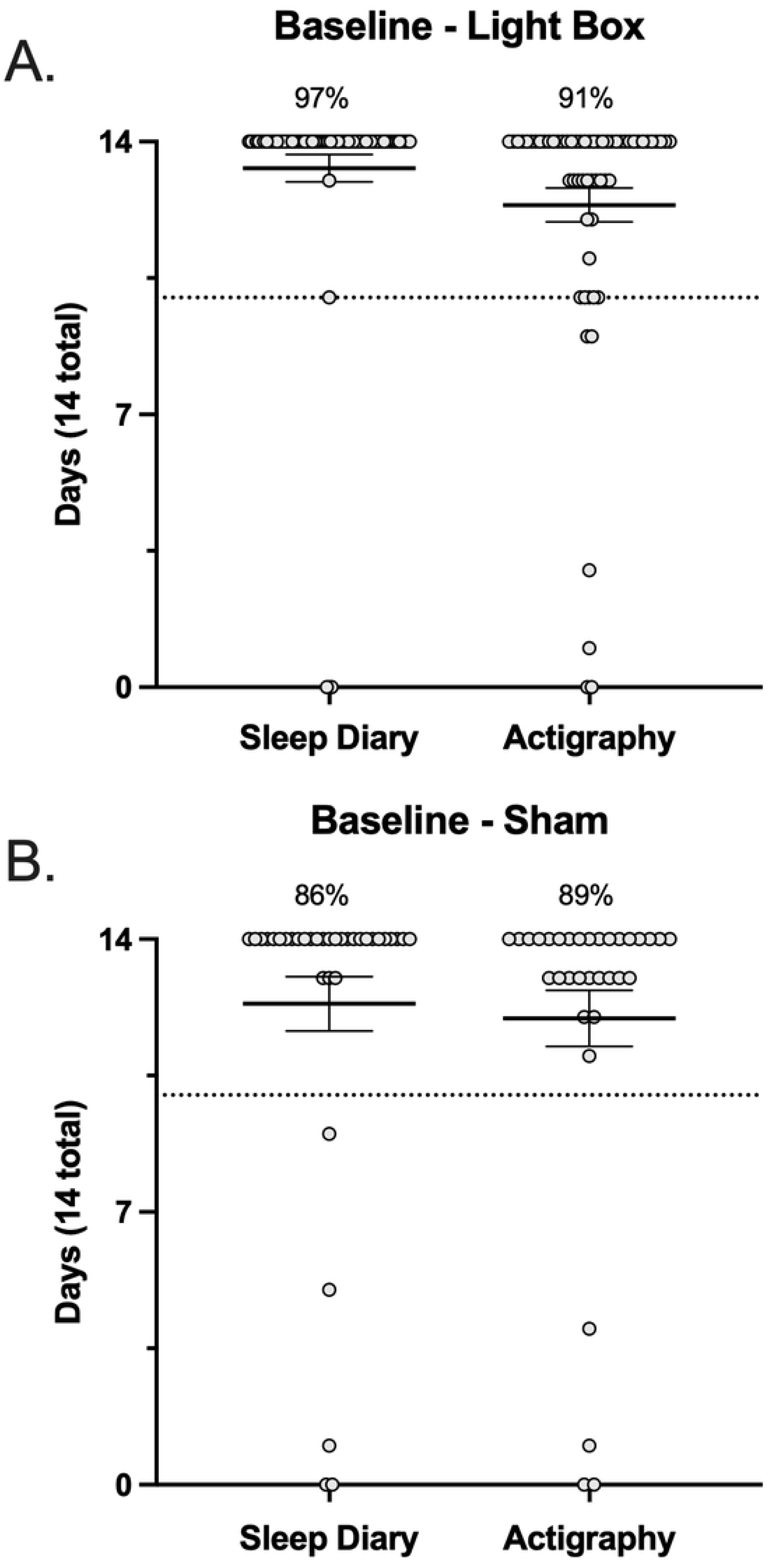
Baseline adherence. Sleep diary and actigraphy data collected at baseline (over 14 days) in the active (**A**) and sham (**B**) conditions. The dashed line indicates the 70% threshold for adherence (10 days). Each data point represents a single participant and corresponds to the number of days they have complete data.

**Figure 5.**
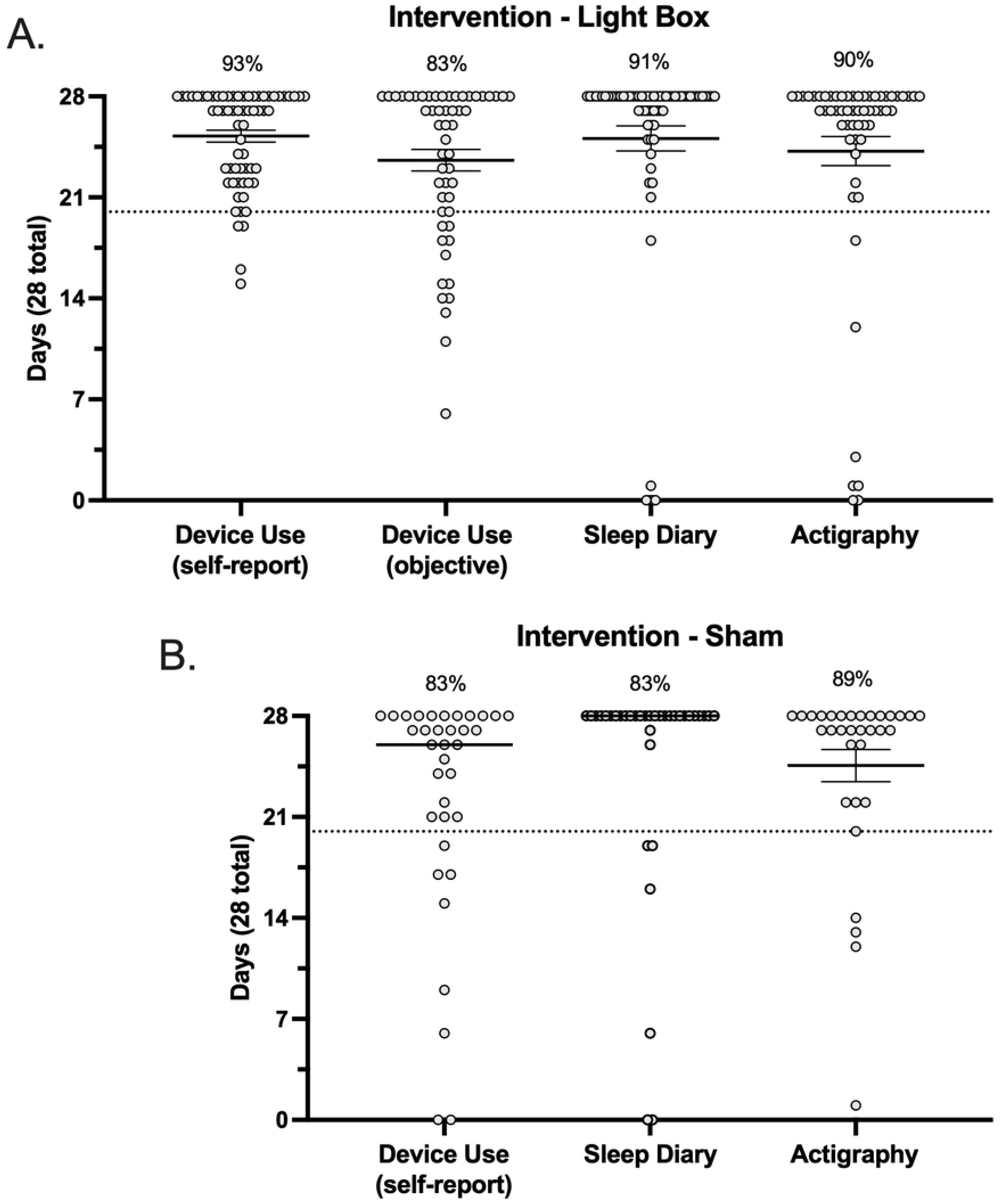
Intervention adherence. Device use (self-report and objective), sleep diary and actigraphy data collected during intervention (over 28 days) in the active (**A**) and sham (**B**) conditions. The dashed line indicates the 70% threshold for adherence (20 days). Each data point represents a single participant and corresponds to the number of days they have complete data.

With respect to our primary outcome (ISI score), >95% of participants in both study arms have provided both pre- and post-intervention data. Supplemental sleep related outcomes included wrist-based actigraphy and sleep diary entries, the completeness for both of which was expressed in terms of the number of days with full/usable data (i.e., out of a maximum of 14 days for the baseline period and out of a maximum of 28 days for the intervention period). For both actigraphy and sleep diaries, we reported >83% of participants have complete data, defined as a minimum of 10 days of data recorded at baseline (**Figure 4A and 4B**) and 20 days of data recorded during the intervention period (**Figure 5A and 5B**), reflecting >70% of days for both periods.

## DISCUSSION

The LION protocol was designed as a sham-controlled, participant-masked randomized controlled trial evaluating effectiveness of MBLT to improve self-reported and objective aspects of sleep quality, with subsequent potential “downstream” effects evaluated in neurobehavioral function, cognition, mood, and quality of life, and blood/sweat based-biomarkers of inflammation in Veterans with mTBI. This project was sponsored by MTBI^2^ and is entering its final year of enrollment and data analysis. At the time of this interim analysis for feasibility and acceptability, we have completed data collection in ∼80% (n=104) of the proposed target sample (*n*=120).

LION was originally conceived as, and initiated in, an in-person format typical for device-based randomized controlled trials. However, in response to the COVID-19 pandemic, this protocol was revised to enable its execution in a fully remote capacity. The remotely executed randomized clinical trial format is innovative in and of itself, but especially so considering this protocol was implemented within the VA. Indeed, success with this current design was the result of many years of preparation in establishing standard operating procedures, integrating VA regulatory, clinical, the Office of Information Technology, information security/privacy, biomedical, mailroom, and other services within the local and national VA system, as well as refinement of the protocol to reduce burden and enhance the participant experience. Specific aspects of this study protocol that were novel and logistically critical to its success include: 1) The implementation of an electronic digital signature service (DocuSign) for obtaining informed consent. This enabled research coordinators to screen, consent, and schedule future visits all on essentially the first point of contact, capitalizing on participant motivation and minimizing the potential for errors or PHI/PII exposure. 2) Establishing the logistical workflow for shipping equipment to and from participants’ place of residence in a manner that minimized participant burden and enabled transfer of biospecimens. 3) Effective utilization of video conferencing technology. 4) Establishing multiple recruitment modalities to effectively reach participants nationwide. 5) Leveraging the digital distribution (via TWILIO™ and REDCap) of key outcome measures (e.g., all participant questionnaire data and sleep diary entries), with automatic data entry directly into REDCap.

The present rate of participant retention (83%) either exceeds or is on par with other randomized clinical trials of similar scale and complexity, e.g., Lam et al: 81% [28]; Youngstedt et al.: 68% [45]. Additionally, these metrics should also be viewed in the context of this protocol being in a fully remote format, which could potentially introduce additional sources of error and/or opportunity for drop out. Accordingly, the present interim analysis refutes this possibility, demonstrating strong protocol feasibility for the remote implementation of morning bright light therapy in Veterans.

Figures 4 and 5 illustrate the spread across primary sleep related outcomes in terms of participants who were <100% adherent in each metric. In each metric there was a small subset of participants who fell below the 70% threshold for adherence. This subset of participants was further explored, and we found that there were no participants who were consistently non-adherent across all metrics, implying that it would not be straightforward to exclude whole participants from analyses, but rather partial exclusion may need to be explored. For example, within the lightbox arm across all 6 metrics (pre/post sleep diary, pre/post actigraphy, and self-report vs objective device adherence), there were n=21 and n=5 participants who were non-adherent in 1 or 2 metrics, and n=1 who was non-adherent in 3 metrics, but zero participants were non-adherent in all metrics. Interestingly, of participants who were non-adherent in only 1 metric, in ∼50% of those participants (n=10) this was with respect to the objective determination of device adherence. In line with the prespecified Intention To Treat analysis, these participants will need to be retained in analyses. Within the sham arm, there were n=9 and n=4 participants who were non-adherent in 1 or 2 metrics, and no participants who were non-adherent in 3, 4, or 5 metrics (note that there was no objective determination of device adherence for the sham condition). There was a relatively even spread across metrics in these participants, i.e., in no single outcome were participants significantly less adherent.

Secondary outcomes in the proportion of participants with complete data were the entire self-reported questionnaire battery, neurocognitive assessment, DLMO, and biospecimen sample. For each of these metrics, all study groups at all time points report ≥75% data completeness defined as 1) no missing questions in the questionnaire battery, 2) all cognitive assessment tests complete, 3) seven DLMO salivettes for each time point, and 4) sweat and/or blood samples for each time point. The majority of outcomes were ≥83% complete, with many at 100%. Only one category, non-TBI sham participants post-intervention, reported a lower than expected rate of biospecimen sampling (60%). This is a small subset of participants (n=10 in total), and thus, likely reflective of normal variability rather than potential bias in study coordinator/participant interaction. In summary, this interim analysis demonstrated that in the current sample of n=104 participants, we have complete data in key outcome measures in >90% of participants, which also aligns with 80-85% of the sample reporting full device compliance. Thus, remotely implementing this randomized controlled trial has not introduced any significant increase in missingness or data incompleteness.

An additional benefit extending from the remote implementation of this trial was our ability to extend recruitment to anyone within the United States. This greatly increased our reach and anecdotally expanded enrollment to participants who would otherwise be unable or unwilling to participate in research. Similarly, nationwide recruitment also conferred a significantly greater degree of racial and ethnic diversity than would otherwise be feasible to accomplish with recruitment confined to the Portland, OR / Vancouver, WA metro area. Indeed, 34.9% of participants identified as female, 23.6% identified as belonging to a racial minority group, and 9.4% identified as Hispanic or Latino. According to the US Census Bureau (2021), 10% (∼1.67 million) of Veterans identify as female, 25.8% (∼4.17 million) identify as an individual from a racial minority group, and 8.6% (∼1.4 million) identify as Hispanic or Latino. Thus, the present trial generally met or exceeded these US Census Bureau defined Veteran demographic statistics. Appreciating that the nationwide Veteran demographic will continue to increase in racial/ethnic diversity over time, we will continue to prioritize recruiting these traditionally under-represented populations.

In conclusion, the present study protocol described the remote implementation of a placebo-controlled participant-masked randomized controlled trial within the VA. Herein we outlined the process by which the remote implementation was achieved and highlight overall study feasibility and acceptability through an interim analysis describing a high degree of participant compliance, device adherence and meeting criteria for providing complete data across all time points. Indeed, remotely implementing a randomized controlled trial imposed significant challenges but this did not compromise data fidelity, integrity, or completeness, as evidenced by the feasibility data reported herein. Data collection is on track to be completed by September 2024 with full efficacy analyses and future directions/applications forthcoming.

## ACKNOWLEDGMENTS

The authors would like to express their sincere appreciation and gratitude for the participation of our research subjects. Additionally, administrative, regulatory, and technical support from Nadir Balba, PhD, Steve Helms, BS, Ryan Opel, BS, Cadence Michel, BS, DC, and Cosette Olivo, BS.

## AUTHOR CONTRIBUTION

1. Conception and design of the study. JEE, EWT, MEO, ADH, TN, BJM, JMG, MML
2. Acquisition and analysis of data. JEE, JSB, ATK, BRL, MDBE, KP, AAM, SJS, JMG, MML
3. Drafting a significant portion of the manuscript or figures (i.e., a substantial contribution beyond copy editing and approval of the final draft, which is expected of all authors). JEE, JSB, MML
4. Sponsorship of study. BAD

## DATA AVAILABILITY STATEMENT

The data underlying this article will be shared on reasonable request to the corresponding author.

